# Genome-wide analysis implicates inner ear development in Ménière’s disease

**DOI:** 10.64898/2026.02.09.26345758

**Authors:** Zhuozheng Shi, Ravi Mandla, Jingjing Li, Xinzhe Li, Zixuan (Eleanor) Zhang, Sixing Chen, Sandra Lapinska, Alexander O. Flynn-Carroll, Bogdan Pasaniuc, Douglas J. Epstein, Iain Mathieson

## Abstract

Ménière’s disease (MD) is a chronic inner ear disorder characterized by recurrent vertigo, fluctuating sensorineural hearing loss, and tinnitus. Despite these distinctive symptoms, its etiology remains poorly understood. We performed a genome-wide meta-analysis of 8,969 cases and 1,962,542 controls across five large biobanks, identifying five independent genome-wide significant loci and estimating an observed-scale SNP heritability of 7% (SE 0.8%), consistent with a modest but significant genetic contribution to MD risk. Fine-mapping and integrative functional analyses implicate two convergent biological processes – developmental regulation of the inner ear, involving *EYA4, EYA1*, and *LMO4* – and retinoic acid metabolism, with loci near *CYP26A1/C1* and *ALDH1A2* suggesting disrupted RA signaling in sensory and fluid-pressure homeostasis. These developmental regulator genes are robustly expressed in fetal and adult human inner ear cell types, supporting a model in which altered developmental programs predispose to adult vestibular and auditory dysfunction. Phenome-wide and genetic correlation analyses further reveal shared genetic architecture between MD and related traits, including vertigo, tinnitus, hearing loss, migraine, and sleep apnea, situating MD within a broader spectrum of sensory and neurological disorders. Collectively, these findings establish a genetic framework for Ménière’s disease risk and implicate developmental regulators and retinoic acid signaling as key contributing pathways.

## Introduction

Ménière’s disease (MD) is a chronic inner ear disorder characterized by debilitating episodes of vertigo, fluctuating sensorineural hearing loss (SNHL), tinnitus, and aural fullness^1–3^. The etiology of MD remains unclear, but it is frequently associated with endolymphatic hydrops, an abnormal accumulation of endolymph within the inner ear^4^. Several pathophysiological mechanisms have been proposed, including fluid imbalance, infection, and vascular dysfunction, but these hypotheses lack direct genetic or molecular support^5^.

MD affects approximately 1 in 2000 individuals, typically presenting in middle to late adulthood, with higher prevalence among women and those of European ancestry^6–10^. Genetic studies indicate a heritable contribution to MD risk, with familial cases (5–15% of patients) following autosomal dominant inheritance with ∼60% penetrance^11^. Rare coding variants in genes including *OTOG, GJD3, FAM136A*, and *DTNA* have been identified in familial MD^12–21^. However, the genetic basis of sporadic MD, which constitutes the majority of cases, remains poorly understood^5^. The contribution of common variants and the proportion of risk they explain have not been systematically defined, leaving the full genetic architecture and downstream molecular mechanisms unresolved.

Recently, large-scale initiatives to expand biobank sample sizes, harmonize phenotypic data, and perform high-quality genome-wide genotyping across diverse populations have transformed the study of complex traits at scale^22–28^. These resources enable well-powered genome-wide association studies (GWAS) and meta-analyses, even for low-prevalence disorders^29–31^. Furthermore, meta-analyses across biobanks allow for the detection of novel risk loci, refinement of heritability estimates, and mapping of genetic variants to molecular function, thereby providing opportunities to address critical gaps in our understanding of the genetic architecture of MD.

We leveraged genome-wide summary statistics from five major biobanks – UK Biobank (UKB), All of Us (AoU), Million Veteran Program (MVP), FinnGen, and Biobank Japan (BBJ) – to characterize the genetic architecture of MD. Through meta-analysis of 8,969 MD cases and 1,962,542 controls from three populations across these biobanks, and downstream fine-mapping and integrative annotation, we identified five independent genome-wide significant signals at three loci, and a SNP-based heritability of 7%. Four of these signals map to potential regulatory elements for *EYA4* and *EYA1*, genes essential for inner ear development. These associations, combined with suggestive evidence at *LMO4*, highlight a developmental regulatory mechanism. Additional signals near *CYP26A1/C1* and *ALDH1A2* implicate retinoic acid signaling, a pathway critical for sensory and fluid-pressure homeostasis. Phenome-wide and genome-wide correlation analyses further demonstrate shared genetic architecture between MD and non-symptom traits, including migraine (r_g_=0.28) and sleep apnea (r_g_=0.20). Collectively, these findings support a polygenic basis for Ménière’s disease driven by regulatory variation in developmental and morphogen pathways, providing a genetic framework for understanding its complex etiology.

## Results

### Meta-analysis identified five independent genome-wide significant variants

We analyzed data across five biobanks – All of Us^24,32^ (n = 383,735; 841 MD cases), Million Veteran Program^27^ (n = 510,058; 2,306 cases), UK Biobank^25^ (n = 420,473; 1,220 cases), FinnGen^26^ (n = 478,519; 3,680 cases), and Biobank Japan^33^ (n = 178,726; 1,290 cases) (**Table 1**). In aggregate, these datasets include 8,969 MD cases and 1,962,542 controls, corresponding to an overall prevalence of 0.46% in our study population. MD status was defined using phecode 386.1 or an equivalent phenotype mapped from ICD codes (H810, 3860, and 38599), with additional quality control steps applied as described in the **Methods**. To assess the contribution of common variants to MD genetic architecture and disease risk, we restricted analyses to variants with minor allele frequency (MAF) > 1%, yielding 13,423,260 variants. MD cases were more frequent in females (**Figure 1a**), and prevalence increased with age **(Figure 1c, 1d**), consistent with prior reports^5^. Across biobanks, 81.4% of MD cases were of European ancestry (**Figure 1b**), and cases were enriched for European ancestry within All of Us (**Supplementary Figure 1**).

**Table 1.** The five GWAS performed on independent datasets, with corresponding country of collection and ancestry filtering of cases and controls, number of cases and controls in each study, and number of SNPs after filtering for minor allele frequency greater than 1%.

| GWAS | Country;<br>Ancestry | Cases/<br>Controls | Case<br>definition | Data Type | Number<br>of SNPs |
| --- | --- | --- | --- | --- | --- |
| All of Us (AoU) | USA; Multi-ancestry | 841/<br>382,894 | Phecode<br>386.1 | Whole Genome<br>Sequenced | 6,795,311 |
| Million Veteran<br>Program<br>(MVP) | USA; European<br>and Admixed<br>American | 2,306/<br>507,752 | Phecode<br>386.1 | Genotyped and<br>Imputed | 9,254,829 |
| UK Biobank<br>(UKB) | UK; European | 1,220/<br>419,253 | UKB<br>phenocode<br>20002, coding<br>1421 | Genotyped and<br>Imputed | 9,549,936 |
| FinnGen | Finland;<br>European | 3,312/<br>474,839 | H81.0 (ICD-10); 3869<br>(ICD-9);<br>38599 (ICD-8) | Genotyped and<br>Imputed | 9,462,981 |
| Biobank Japan<br>(BBJ) | Japan; East<br>Asian | 1,290/<br>177,436 | Phecode<br>386.1 | Genotyped and<br>Imputed | 7,440,824 |

**Figure 1.**
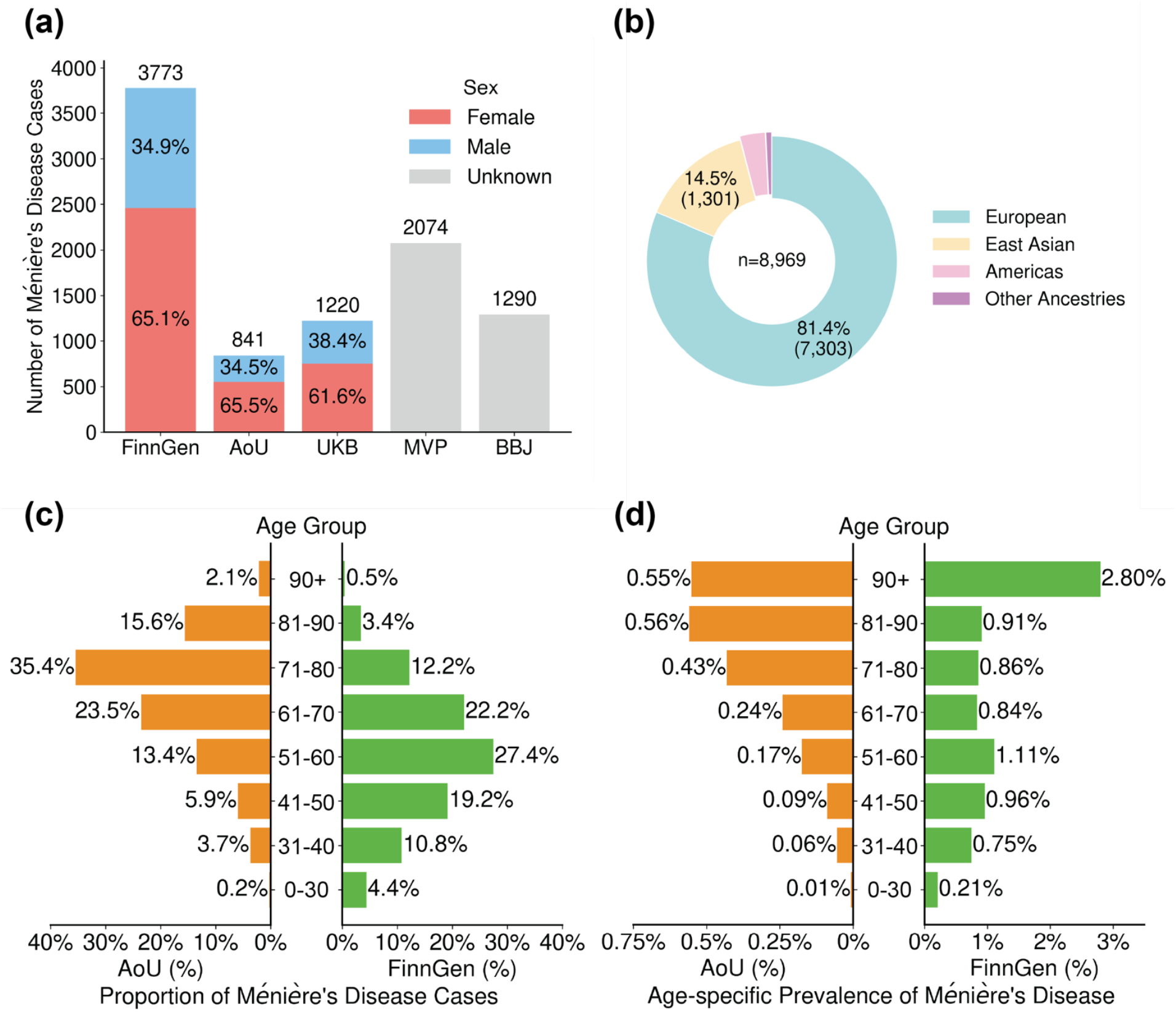
Demographics of Ménière’s Disease Cases. **a.** Sex distribution of Ménière’s Disease cases across FinnGen, All of Us, and UK Biobank. Information on the Million Veteran Program and Biobank Japan was not available. **b**. Genetically inferred ancestry distribution of MD cases across five contributing studies (**Methods**). Other genetically inferred ancestries include African, South Asian, and Middle Eastern. **c**. Age distribution of Ménière’s disease cases in All of Us and FinnGen, categorized by age bins, in proportion. Information on the Million Veteran Program, the UK Biobank, and Biobank Japan was unavailable. **d**. Age-specific prevalence of Ménière’s disease cases in All of Us and FinnGen.

We conducted a GWAS in All of Us using Regenie, and then performed a fixed-effects meta-analysis with summary statistics from the four other studies^24–27,33^ using the inverse-variance weighted scheme in METAL^34^. For each study, we filtered out variants with minor allele frequency <0.01 or minor allele count <10. We estimated SNP-based heritability to be 7% (SE=0.8%) on the observed scale using LD Score Regression^35^ (LDSC) applied to the meta-analyzed summary statistics. Minimal genomic inflation (λ_gc_ = 1.01) in the meta-analyzed result indicated negligible confounding before correction. We identified five independent genome-wide significant index SNPs (P<5×10^-8^; **Figure 2; Table 2)**, defined as the most significant variant within a 1 Mb window, clumped at r^2^ > 0.05, and supported by at least one additional genome-wide significant SNP (**Supplementary Table 1**). Three other loci were just below the significance threshold (**Methods;** 5×10^-8^ < P < 5×10^-7^), and we highlight signals near *LMO4* and *ALDH1A2* due to their biological relevance (**Supplementary Table 2, 3**).

**Table 2.**
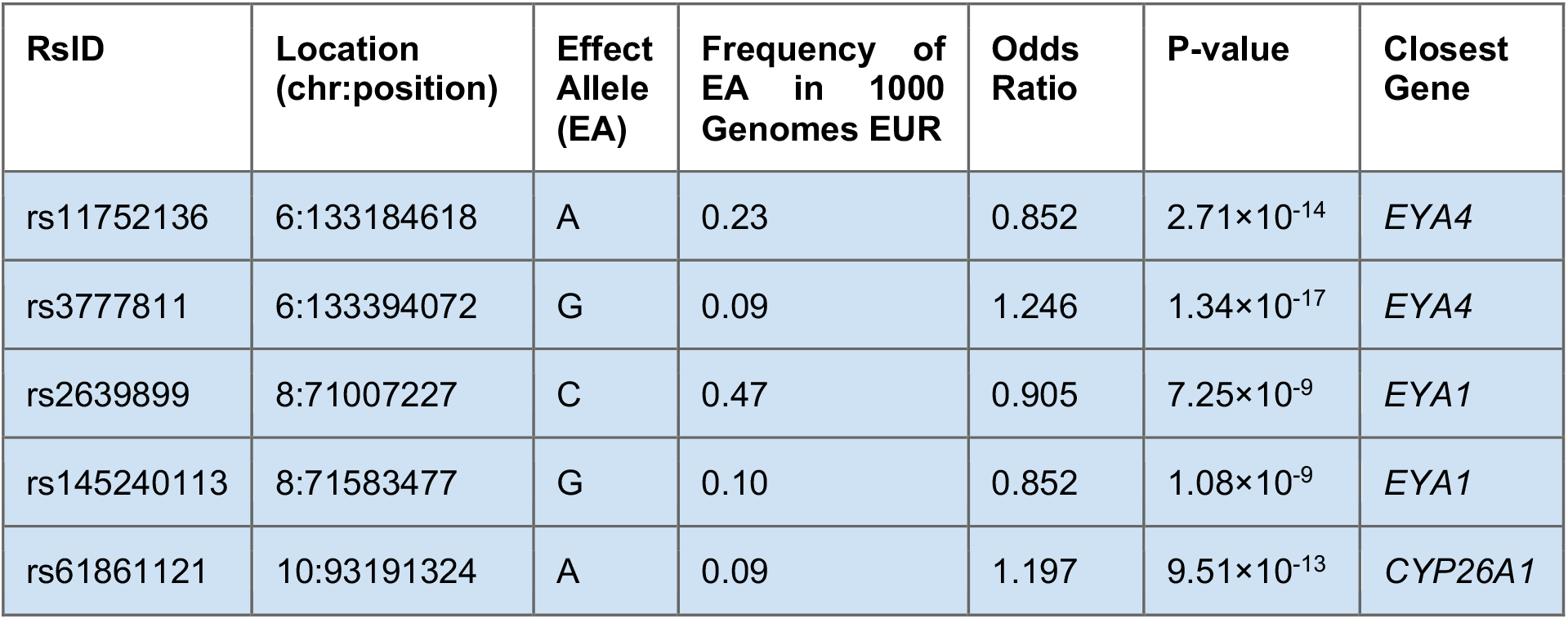
The five genome-wide significant index SNP associations, with corresponding RsID, location in hg38, effect allele, frequency of effect allele in 1000 Genomes EUR population, odds ratio, P-value, and closest gene by distance.

**Figure 2.**
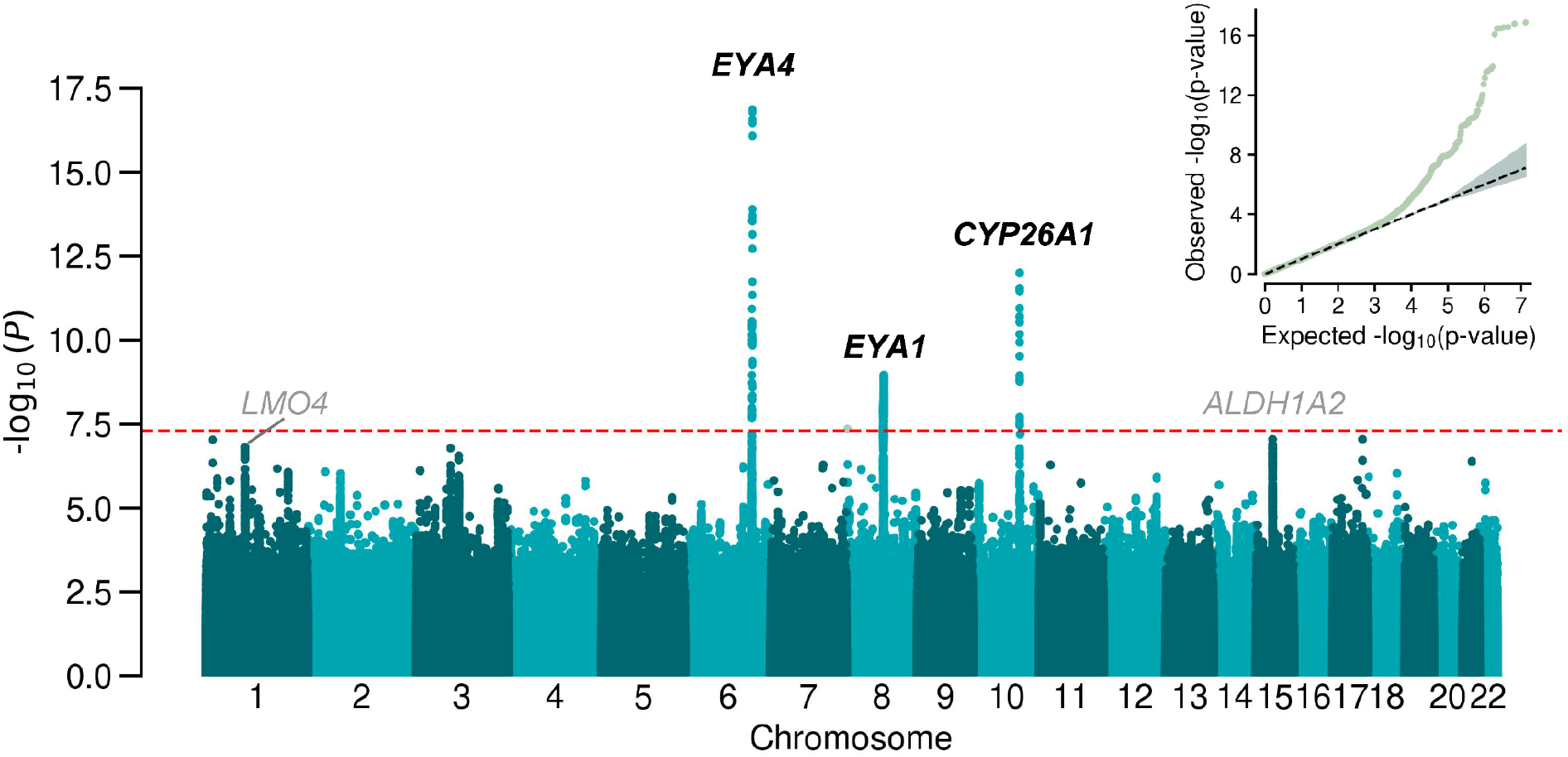
Manhattan plot of meta-analysis. GC-corrected -log10 of P values from the meta-analysis of multi-ancestry Ménière’s Disease GWASs. The red dashed line indicates the genome-wide significance threshold of 5e-8. Genome-wide significant variants, where no other significant variant is within 1 Mb, are plotted in grey. The nearest genes to the index SNPs are annotated in black. Additional loci discussed in the text are annotated in grey. Inset: quantile-quantile plot of the meta-analysis with genomic control applied.

### Two independent signals at *EYA4* suggest a developmental contribution

We next examined the two independent association signals located 209 kb apart at *EYA4* (index SNPs rs11752136 and rs3777811, *r*^*2*^ = 0.003; **Table 2; Figure 3a**). *EYA4* encodes a member of the evolutionarily conserved eyes absent (EYA) protein family and functions as both a transcriptional coactivator and a phosphatase. EYA4 has essential roles in auditory development and function and loss-of-function mutations lead to autosomal dominant sensorineural hearing loss (DFNA10)^36–40^. Significant SNPs clumped with rs11752136 overlap candidate cis-regulatory elements located ∼60kb upstream of *EYA4* (Encode track, UCSC browser), the *EYA4* transcription start site (TSS), the first intron, and an enhancer in intron 1. rs3777811 is located 155 kb downstream of the *EYA4* TSS in intron 3 (**Figure 3a**). Fine-mapping with SuSiE^41^ resolved the region into two credible sets corresponding to the two index SNPs; however, high LD precluded confident identification of a single causal variant in either region (**Figure 3b; Supplementary Table 4-6**). *EYA4* shows a strong gene-level association with MD (MAGMA P=3.3×10^-10^; **Supplementary Table 10**).

**Figure 3.**
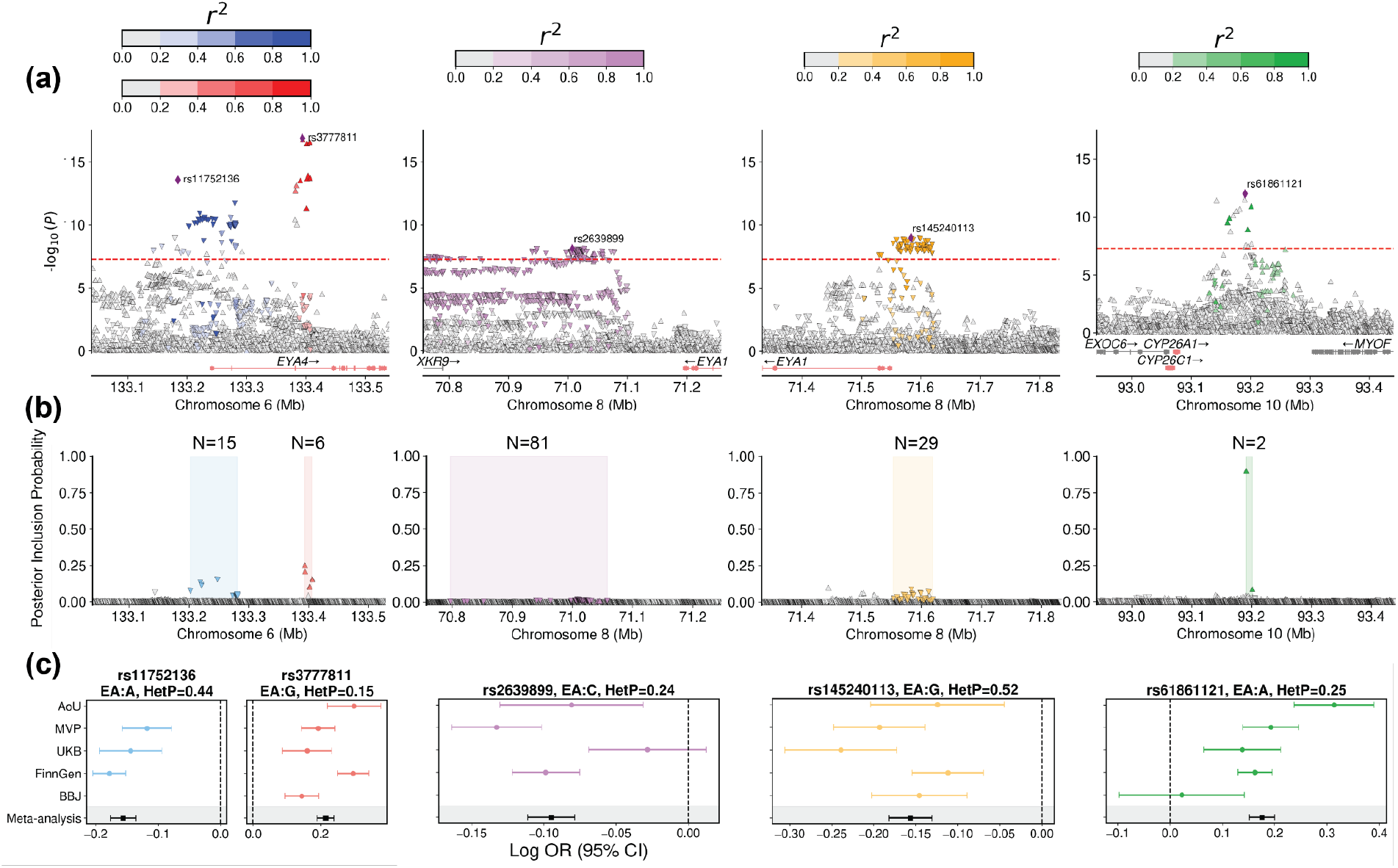
Characterization of genome-wide significant loci for Ménière’s disease. **(a)** -log_10_ P-values from the meta-analysis plotted against genomic positions for each index SNP region. The window size of the plots is 500kb. Genes of interest are colored in red along the x-axis. Red dotted line denotes genome-wide significance (P<5×10^-8^). (**b**) Posterior inclusion probabilities (PIPs) from SuSiE fine-mapping plotted against genomic position for each index SNP region. Shading denotes the start and end of credible sets, and the number of SNPs in the credible set is annotated. (**c**) Forest plots of odds ratios as OR for each GWAS study and meta-analysis with 95% confidence intervals on the x-axis for each index SNP. For each index SNP, effective allele and heterogeneity P-values are included in the title.

*EYA4* is expressed in sensory and non-sensory cell types of the cochlea and vestibular system at fetal and adult stages according to single-cell RNA-seq data from the human inner ear (**Figure 4**). Although eQTL data are not currently available for the inner ear, rs3777811 is an eQTL for *EYA4* in several brain tissues, with the MD risk allele associated with increased *EYA4* expression **(Supplementary Table 11)**, whereas weak LD proxies of rs11752136 (rs971589; r^2^ = 0.48) are eQTLs for *EYA4* across brain, heart, and tibial nerve tissues **(Supplementary Table 11)**. Phenome-wide analysis showed that rs3777811 is associated with increased risk of sensorineural hearing loss (SNHL), tinnitus, and general ear disorders, core symptoms that are genetically correlated with MD (**Figure 5a, b**), as well as decreased risk of sleep apnea (**Supplementary Table 16**). Notably, rs3777811 is distinct from, and not in LD with, the previously identified coding variant rs9493627, which is associated with SNHL^42^. In contrast, rs11752136 is an MD-specific signal and shows no association with related phenotypes (**Supplementary Table 15**). Together, these complementary signals suggest distinct regulatory mechanisms, one shared with SNHL and one specific to MD. Functional characterization of these two variants may help identify molecular mechanisms that are both specific to MD and shared with related disorders.

**Figure 4.**
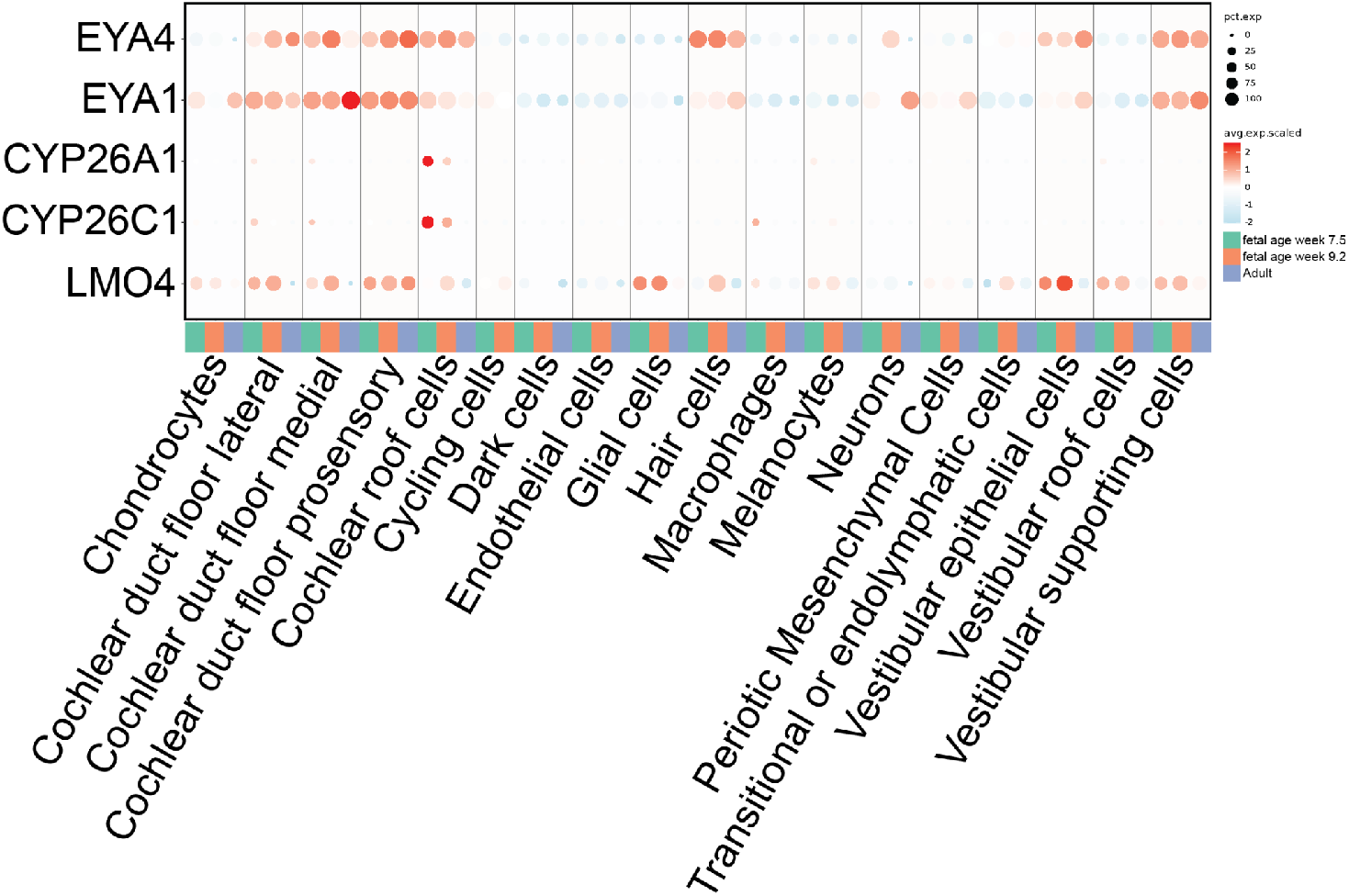
Dotplot of single-cell expression from human inner ear scRNAseq across developmental stages. Dot plot shows the expression of EYA4, EYA1, CYP26A1, CYP26C1, and LMO4 across diverse human inner ear cell types at three developmental stages (fetal week 7.5, fetal week 9.2, and adult). Dot size represents the percentage of expressing cells, and color indicates scaled average expression. Data were obtained from the human inner ear atlas^60^ via the gEAR Dashboard^61^.

**Figure 5.**
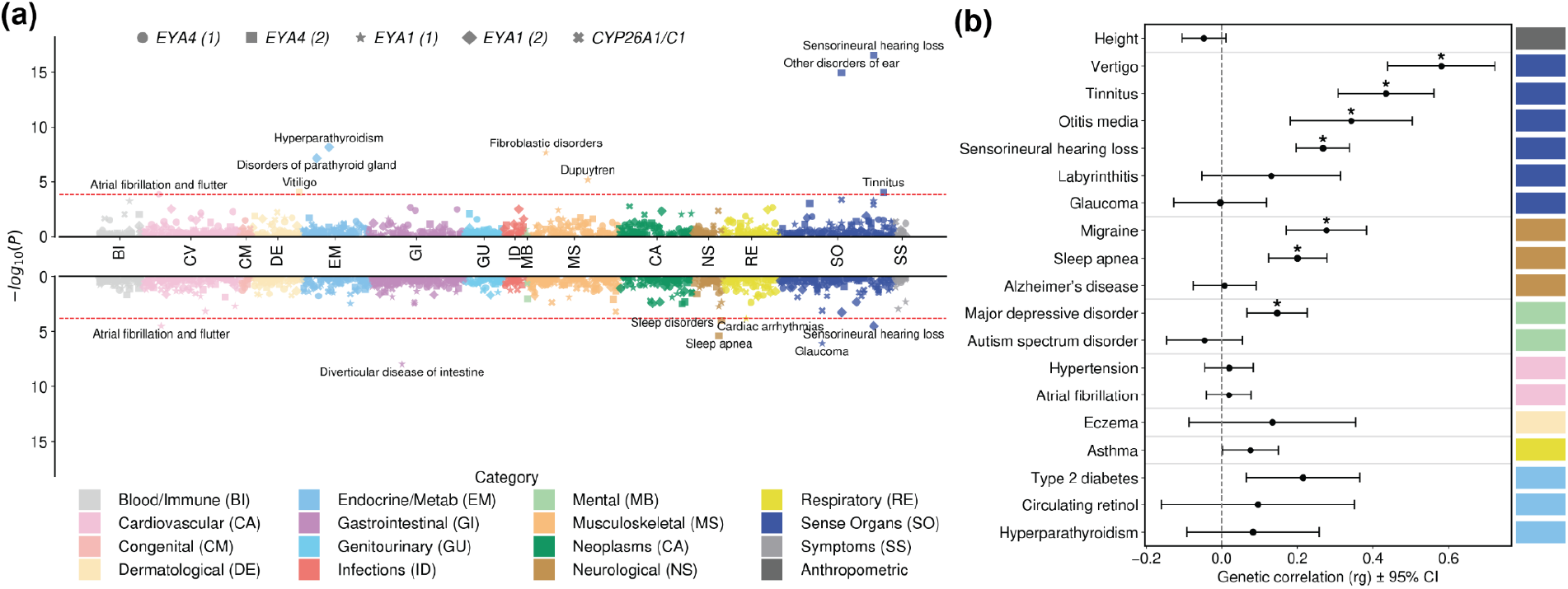
Phenotypic characterization. **(a)** Phenome-wide associations from harmonized FinnGen, MVP, and UKB GWAS results of the five index SNPs across disease categories are plotted. The red dotted line denotes the Bonferroni-corrected P-value threshold. The top plot indicates the same direction of effect as Ménière’s disease, while the opposite direction is indicated in the bottom plot. Phenotypes with significant association are denoted. **(b)** Forest plot of genetic correlation estimates of Ménière’s disease between phenotypes across categories. Asterisk denotes Bonferroni-corrected significance.

### Two independent signals at *EYA1* further emphasize the role of development

At *EYA1*, we found another two independent index SNPs, rs2639899 and rs145240113 (r^2^= 1×10^-4^; **Table 2; Figure 3a**). rs2639899 lies 192 kb downstream of *EYA1* in a candidate cis-regulatory element within a genomic region on chromosome 8 that also features a cluster of five ultraconserved non-coding elements (UCNEBase track, UCSC Genome Browser). rs145240113 is located 35 kb upstream of the *EYA1* TSS. Fine-mapping delineated two credible sets corresponding to these index SNPs, though again, extensive local LD precluded identification of causal variants (**Figure 3a, 3b; Supplementary Table 4, 7, 8**).

*EYA1*’s association with MD reached gene-level significance (MAGMA P=1.3×10^-5^; **Supplementary Table 10**), and similar to *EYA4*, is expressed in the human inner ear at fetal and adult stages (**Figure 4**). *EYA1* encodes a homologous dual-functioning transcriptional regulator and phosphatase, and its haploinsufficiency causes Branchio-Oto-Renal (BOR) syndrome, a congenital disorder characterized by malformations of the outer, middle and inner ear, hearing loss, and renal anomalies^43–45^. Although EYA1 and EYA4 share similar phosphatase substrates^36^, EYA1 functions earlier in otic development than EYA4, with EYA1-deficient ears arresting at the otic vesicle stage^46^.

rs2639899 is an eQTL for *EYA1* in esophageal mucosa. Consistent with the effect of rs3777811 on *EYA4*, the MD risk allele of rs2639899 is associated with increased *EYA1* expression (**Supplementary Table 12)**. Besides Ménière’s disease, the risk-increasing allele of rs2639899 is associated with several phenotypes, including reduced risk of glaucoma, while the risk-increasing allele of rs145240113 is associated with increased risk of hyperparathyroidism and decreased risk of SNHL (**Figure 5a; Supplementary Table 17, 18**).

### An association at *LMO4* is consistent with its developmental role in mice

We observed a suggestive association at rs1199565, located 101 kb downstream of *LMO4* (P = 1.46×10^-7^; **Supplementary Table 2; Supplementary Figure 2**) and annotated as an eQTL in subcutaneous adipose tissue (**Supplementary Table 13**). Although below genome-wide significance, this signal is noteworthy given the established role of *LMO4* in vestibular and cochlear morphogenesis: loss of *LMO4* expression in mice disrupts inner-ear development and impairs balance^47,48^. In the human fetal inner ear, single-cell RNA-seq data show strong *LMO4* expression in vestibular epithelial and cochlear duct floor cells (**Figure 4**), suggesting a conserved developmental function. Together with the *EYA4* and *EYA1* loci, this locus reinforces a broader theme in which dysregulation of genes guiding inner-ear formation contributes to Ménière’s disease susceptibility.

### The *CYP26A1/C1* locus implicates retinoic acid signaling

On chromosome 10, the index SNP rs61861121 lies downstream of *CYP26A1, CYP26C1, EXOC6, and MYOF* (**Table 2; Figure 3a, 3b**). Fine-mapping identified rs61861121 as a causal SNP within a two-variant 95% credible set, with a posterior inclusion probability of 0.90 (**Figure 3b; Supplementary Table 4, 9**). rs61861121 is not an eQTL for any nearby genes, and none reached gene-level significance in MAGMA (*CYP26A1* P = 0.03; *CYP26C1* P = 0.05, *EXOC6* P=0.12, *MYOF* P=0.05; **Supplementary Table 10**), leaving the causal gene unresolved. Nonetheless, functional considerations suggest that *CYP26A1/C1* are plausible candidates. Both genes encode enzymes that degrade retinoic acid to maintain spatial gradients during embryogenesis, a process essential for patterning of the retina, hindbrain, and otic placode^49–52^. The functional paralog of *CYP26A1/C1 in* mice – *CYP26B1* – has a critical role in vestibular sensory epithelia formation by shaping retinoic acid availability in the embryonic inner ear^53^. The vestibular structures affected are responsible for detecting head motion, directly relevant to MD. Although rs61861121 shows no association with other phenotypes after Bonferroni correction (**Figure 5a**), a more lenient threshold (**Methods**) suggested associations with multiple eye and ear phenotypes, including glaucoma, peripheral retinal degeneration, and sensorineural hearing loss (**Supplementary Table 19**). This convergence suggests a role for retinoic acid signaling in a broader category of fluid tension disorders spanning glaucoma, intracranial hypertension, and MD^54^. The potential role for retinoic acid signaling is further supported by a suggestive association at *ALDH1A2* (rs6493979; P = 8.6×10^-8^; **Supplementary Table 2; Supplementary Figure 2**), which is MAGMA significant (P = 3.07×10^-7^; **Supplementary Table 10**) and has moderate expression in vestibular epithelial cells (**Supplementary Figure 3**). *ALDH1A2 is* directly involved in retinoic acid synthesis^55–58^ and associated with cerebral ventricle volume^59^, which, again, points to disruption of retinoic acid metabolism in MD etiology and a link with other fluid tension disorders.

## Discussion

In this study, we report the first large-scale genome-wide association study of Ménière’s disease (MD), integrating data from nearly two million individuals across five major biobanks. An observed-scale SNP heritability of 0.07 indicates a measurable but modest contribution of common variants to disease risk. Common variation in genes associated with familial MD does not appear to make a substantial contribution to sporadic cases. We identify five genome-wide significant loci with functional and phenotypic evidence implicating developmental regulation of the inner ear, and retinoic acid–mediated signaling. These findings expand our understanding of the genetic basis of MD beyond familial rare variants and point to biological pathways important in disease.

The most robust associations map to *EYA4* and *EYA1*, transcriptional regulators essential for otic placode and inner ear development^38–40,44–46^. Both loci harbor two independent non-coding signals. Two of the four index SNPs – rs11752136 at *EYA4* and rs145240113 at *EYA1* – act as eQTLs in various tissues, and MD risk alleles of both are associated with increased expression of *EYA4* and *EYA1* (**Supplementary Table 11, 12**). Meanwhile, a significant association, rs3836957, located at *EYA4* and clumped with rs11752136, was identified as an enhancer (**Supplementary Table 20**). These patterns suggest that MD-associated alleles modulate gene expression rather than causing loss of function. Expression analysis of human inner ear single-cell RNA-seq data further supports the relevance of these genes: both *EYA4* and *EYA1* are expressed in vestibular and cochlear epithelial cell types during fetal and adult stages, while *LMO4* is expressed in vestibular and cochlear duct floor cells (**Figure 4**). These findings converge on developmental dysregulation of sensory epithelia as a shared mechanism linking MD to congenital and acquired hearing disorders. Consistent with this model, *EYA4* variants have been implicated in autosomal dominant sensorineural hearing loss^36,37,40^, *EYA1* is causal in Branchio–Oto–Renal syndrome^45^, and loss of *LMO4* expression leads to maldevelopment of the mouse inner ear^43,44,47,48^. The extension of these developmental pathways to MD, typically a late-onset, sporadic condition^2,5^, suggests that subtle regulatory perturbations in the expression of developmental genes can predispose to adult vestibular and auditory dysfunction, in agreement with a recently proposed model of MD pathophysiology^62^.

A complementary biological theme arises from the association near *CYP26A1/C1* and the suggestive signal at *ALDH1A2*. These genes encode enzymes that respectively degrade and synthesize retinoic acid (RA), maintaining local RA gradients essential for hindbrain and otic patterning^50–52,63–66^. Fine-mapping highlights rs61861121 as a likely causal variant with high posterior probability, pointing to regulatory perturbation of *CYP26A1/C1* rather than protein-altering effects. In mice, the paralog *Cyp26b1* controls vestibular sensory epithelium formation through spatial RA regulation, directly linking this pathway to balance^53^. These associations align with prior hypotheses connecting RA metabolism to endolymphatic homeostasis and provide a developmental-to-physiological bridge for MD pathogenesis^62,63^.

Phenome-wide association analyses and genome-wide genetic correlations highlight the intersection between MD and related sensory or neurological traits. The strongest polygenic overlaps are observed with vertigo, tinnitus, and sensorineural hearing loss, which are core symptoms of MD. We also observe a positive genetic correlation with migraine, a comorbidity of MD that may share vascular or neurogenic mechanisms^67^. However, despite a positive genetic correlation, GWAS signals for vertigo and migraine do not overlap with MD^68,69^, indicating that the largest effects are not shared. We also find no evidence that common variants at genes associated with familial MD contribute to risk of sporadic MD. On the other hand, we find overlap in GWAS signals but low genetic correlations for fluid-tension related traits, including glaucoma, suggesting a shared biological pathway, albeit one that only explains a small proportion of risk.

Together, our results establish MD as a polygenic disorder influenced by regulatory variation in genes governing inner ear development and retinoic acid signaling. These factors only explain a small fraction of risk, and it remains to be seen how they interact with other genetic factors or environmental triggers. These findings lay the groundwork for future functional studies using human inner ear organoids and in vivo models to test how subtle dysregulation of pathways involving *EYA1/4* and *CYP26A1/C1* pathways affects sensory epithelia formation and fluid balance. Beyond mechanistic insight, this work provides a foundation for constructing polygenic risk models and exploring comorbidity across sensory and neurological disorders. Expanding these analyses to larger and more ancestrally diverse cohorts and integrating inner ear–specific regulatory eQTL data will be the next steps toward a comprehensive understanding of MD genetics and pathology and its place within the broader landscape of auditory and vestibular disease.

## Methods

### Contributing Biobanks and Genome-wide Association Studies (GWAS)

#### All of Us Research Program

The *All of Us Research Program (AoU)* is a US-based biobank with genomics data, electronic health record (EHR), and survey information from diverse populations^24^. We performed a GWAS of Ménière’s disease using Regenie^31^, including age, sex, age2, age2*sex, and the top 10 PCs as covariates. We obtained genotypes from short-read whole-genome sequencing in the Curated Data Repository (CDR) v8 release, and defined phenotypes using phecode 386.1. We filtered variants by population-specific allele frequency > 1% or allele count > 100 in any ancestry subgroup. We performed additional quality control in PLINK 2 using --maf 0.01, --geno 0.02, and --mind 0.05 across all individuals. Related individuals were removed. We defined cases as participants with ≥ 2 phecode encounters, and controls as those with none.

#### Million Veteran Program

The Million Veteran Program (MVP) is a large and ancestrally diverse U.S. biobank linking genomic data to detailed EHR and survey information^70^. We used summary statistics done by MVP from the GIA gwPheWAS release^27^, where the Ménière’s disease GWAS (phecode 386.1) was performed with SAIGE^71^, among European and Admixed American participants (2,306 cases, 507,752 controls). Cases were defined as participants with ≥ 2 phecode encounters, and controls as those with none. Ancestry was inferred via PCA-based random-forest classifiers. Genotypes were imputed using a hybrid reference panel of African Genome Resources + 1000 Genomes Phase 3 v5 and filtered by imputation quality > 0.3, minor allele count > 20, call rate > 0.975 for MAF > 0.01, and > 0.99 for MAF < 0.01^27^.

#### UK Biobank

*The UK Biobank (UKB)* is a prospective cohort of ∼500,000 participants with genotype and EHR data from the United Kingdom^23^. We used the *Pan-UK Biobank* European-ancestry GWAS (phenocode 20002, coding 1421, SAIGE model)^25^, which included 1,220 cases and 419,253 controls. Genetic ancestry was inferred using PCA-based random-forest classifiers, and only unrelated individuals (PC-Relate) were included. Genotypes were imputed to the UKB version 3 panel (∼97 million variants) and filtered by INFO > 0.8 and minor allele count > 20^25^.

#### FinnGen

*FinnGen* is a nationwide research initiative combining Finnish biobank data with health registry records^26^. We used GWAS summary statistics from the R12 release for endpoint H8_Ménière, analyzed with Regenie. The study included 3,312 cases and 474,839 controls of European ancestry. Phenotypes were harmonized across ICD-10 (H810), ICD-9 (3860), and ICD-8 (38599) codes. Genotypes were imputed to the population-specific SISu v4.2 reference panel and filtered by INFO > 0.9 and MAF > 0.01.

#### Biobank Japan

*Biobank Japan* is a large hospital-based biobank that links genomic data with clinical information from ∼200,000 Japanese participants across phenotypes^28,72^. The BBJ GWAS was conducted using SAIGE to test for associations between ∼13 million imputed variants (imputed to the 1000 Genomes Phase 3 East Asian panel) and clinically ascertained diagnoses. For Ménière’s disease, the analysis included 1,290 cases and 177,436 controls, defined by phecode 386.1. Quality control followed the BBJ protocol^72^: variants with MAF < 0.01, imputation INFO < 0.8, or Hardy–Weinberg P < 1e-6 were excluded, and participants with call rate < 0.98 or excess heterozygosity were removed. Association testing adjusted for age, sex, top 20 genetic PCs, and genotyping batch.

### Ancestry Definition

Genetic ancestry assignments for All of Us, the Million Veteran Program, and UK Biobank were obtained from each cohort, where ancestry inference was performed using principal component analysis (PCA) and random-forest classifiers trained on the Human Genome Diversity Project and the 1000 Genomes Project^24,25,27^. In FinnGen and Biobank Japan, participants were recruited from genetically homogeneous Finnish and Japanese populations, corresponding broadly to EUR and EAS ancestries, respectively.

### GWAS Meta-analysis

We conducted the meta-analysis using METAL with summary statistics from five GWAS described above^24–27,34,72^. We lifted over Pan-UKB and BBJ summary statistics from GRCh37 to GRCh38 using the UCSC liftOver tool^73^. We kept single study-private variants, and removed variants with minor allele frequency <0.01 or with minor allele count <10. We applied genomic control to all datasets before meta-analysis and again on the meta-analyzed result. We used the STDERR scheme to perform meta-analysis, utilizing weighted effect size estimates and inverted standard errors. We calculated the effective sample size to replace the overall sample size^34^. Cochran’s Q and its P-value as measures of heterogeneity were calculated by METAL. We had 14,096,168 SNPs after meta-analysis, and kept a total of 13,423,260 that are also in the 1000 Genomes high-coverage Phase 3 reference panel^74^ for downstream analyses.

### Heritability Estimate

We estimated SNP-based heritability using LD Score Regression^35^ (LDSC). Meta-analysis summary statistics for Ménière’s disease (MD) were first processed using the munge_sumstats.py script provided with LDSC. We excluded strand-ambiguous SNPs and aligned all variants to the 1000 Genome Phase 3 high-coverage reference genome^74^. For LD reference, we computed LD scores using 632 European populations from the 1000 Genomes Project Phase 3 high-coverage data.

We estimated heritability on the observed scale, assuming a case-control design with 8,969 MD cases and 1,962,542 controls. We also attempted, but failed, to convert observed-scale heritability to liability-scale heritability using an assumed population prevalence of 0.2% and the methods of Lee et al. and Ojavee et al.^75,76^

### Index SNP selection

We defined Genome-wide significant hits at P<5e-8 and suggestive associations at P<5e-7. We obtained index SNPs, or index hits, through LD-clumping using Plink 1.9, with –clump-r2 of 0.05 and – clump-kb of 1 million bps^77^. Genotype data of 632 European individuals from the 1000 Genomes Project Phase 3 high-coverage variants data were used as reference panels^74^. The clumping threshold for P-value is first set to 5e-8 and then loosened to 5e-7 for regions previously having no clumped result. Loci with P values below the thresholds but clumped with no other variants were defined as significant or suggestive loci without support.

### Variant Consequences

To predict the consequences of the variants, we employed Ensembl Variant Effect Predictor (VEP)^78^. The input to VEP was the list of SNPs that are significant and clumped with each index SNP.

### Fine-Mapping

To localize putative causal variants within genome-wide significant regions, we performed statistical fine-mapping using the Sum of Single Effects (SuSiE)^41^. The fine-mapping window was ±500 Kb centered on each index SNP, and the midpoint of two index SNPs on chromosome 6. For each region, SNPs that are present across the five contributing biobanks were kept. We constructed an ancestry-weighted LD panel by effective sample size in the meta-analyzed GWAS from the 1000 Genomes Project Phase 3 high-coverage data^74^. We confirmed accurate LD matching between the reference and meta-analysis data by assessing LD–Z concordance (s ≤ 0.003 across all loci). For each locus, SuSiE was run with up to 10 potential causal components (L = 10) and default prior variance settings. 95% credible sets (CS) were defined as the minimal subset of variants whose cumulative posterior inclusion probabilities (PIP) reached 0.95.

### PheWAS of Index SNPs

To assess the phenotypic pleiotropy of lead SNPs from the MD GWAS meta-analysis, we queried publicly available PheWAS results from the FinnGen-MVP-UK Biobank meta-analysis, accessed via FinnGen Data Freeze 12^26^. For each index SNP, we extracted its association statistics across a curated panel of 330 harmonized phenotypes. Only associations reaching Bonferroni significance of 1.5e-4 were considered statistically significant.

### eQTL Characterization

To assess regulatory relevance, we checked whether each index SNP was a significant cis-eQTL in any tissue based on GTEx v10 single-tissue cis-eQTL summary statistics^79^. We considered variant-gene pairs passing permutation-based significance thresholds (FDR < 0.05) across 50 tissues.

### MAGMA Gene Association

We performed gene-level association analysis using MAGMA to map SNP-level GWAS meta-analysis summary statistics to gene-level signals, where individual SNP p-values were combined into gene test-statistics^80^. We mapped SNPs in meta-analysis summary statistics to genes through annotation using a protein-coding gene location file in build 38 obtained from the NCBI site, including transcription start site and end site (https://cncr.nl/research/magma/). We adopted an annotation window of 10kb before the transcription start site and 10kb after the end site to include SNPs around the gene. Then, genes were prioritized using the gene annotation file and the meta-analysis summary statistic, where genotype data from the European population in the phase 3 1000 genomes project served as reference data, and a uniform sample size was specified^74^. We identified significant gene associations after Bonferroni correction using the number of annotated genes.

### Human Inner Ear Gene Expression

Single-cell RNA-sequencing data were obtained from the human inner ear atlas^60^ and accessed through the gEAR Dashboard (https://umgear.org/). Processed gene expression matrices and cell-type annotations provided by the atlas were used for downstream visualization. All analyses were performed in R using the Seurat package (version 5.3.1). Gene expression values were normalized and scaled using Seurat’s default normalization and scaling workflows. Dot plots were generated using the Seurat function *DotPlot*.

### Genetic Correlation

We estimated genetic correlations using LD Score Regression^35^ (LDSC). Data processing and LD score calculation were discussed in the section on heritability estimate. To assess shared genetic architecture between MD and other traits, we performed pairwise genetic correlation analysis using the ldsc.py --rg command. We processed summary statistics for 18 external GWAS summary statistics using the same pipeline, and estimated genetic correlations using the same LD reference panel. For the list of external GWAS, please see **Supplementary Table 21**.

## Data Availability

Summary statistics for the meta-analysis have been deposited in the GWAS catalog, accession number GCST90809428.

## Acknowledgments

We gratefully acknowledge All of Us participants for their contributions, without whom this research would not have been possible. We also thank the National Institutes of Health’s All of Us Research Program for making available the participant data examined in this study. We also want to acknowledge the participants and investigators of the FinnGen study

This work was supported by the National Institute of Aging R01AG085518 (B.P), the National Institute of Mental Health R01MH115676 (B.P), the National Institute of General Medical Sciences R35GM133708 (I.M), and the National Institute on Deafness and Other Communication Disorders R01DC021475 (D.J.E). The content is solely the responsibility of the authors and does not necessarily represent the official views of the National Institutes of Health.

## Contributions

I.M. and D.J.E. conceived the project. I.M., B.P., D.J.E., and Z.S. designed experiments. Z.S. performed experiments and statistical analyses with assistance from R.M., X.Z., Z.Z., S.L., and A.F.C. J.L. QCed the genomic data in the All of Us Research Program. S.C. generated Figure 4. I.M., D.J.E, and B.P. supervised the project. Z.S., I.M., D.J.E, and B.P. wrote the paper with feedback from all authors.

## Declaration of generative AI and AI-assisted technologies in the writing process

During the preparation of this work, ChatGPT was used in order to improve grammar and readability in the method section. After using this tool/service, the authors reviewed and edited the content as needed and take full responsibility for the content of the publication.

## References

1. Minor, L. B., Schessel, D. A. & Carey, J. P. Ménière’s disease. Curr. Opin. Neurol. 17, 9 (2004).

2. Sajjadi, H. & Paparella, M. M. Meniere’s disease. The Lancet 372, 406–414 (2008).

3. Pyykkö, I., Nakashima, T., Yoshida, T., Zou, J. & Naganawa, S. Ménière’s disease: a reappraisal supported by a variable latency of symptoms and the MRI visualisation of endolymphatic hydrops. BMJ Open 3, e001555 (2013).

4. Merchant, S. N., Adams, J. C. & Nadol, J. B. J. Pathophysiology of Ménière’s Syndrome: Are Symptoms Caused by Endolymphatic Hydrops? Otol. Neurotol. 26, 74 (2005).

5. Nakashima, T. et al. Meniere’s disease. Nat. Rev. Dis. Primer 2, 1–18 (2016).

6. Tyrrell, J. S., Whinney, D. J. D., Ukoumunne, O. C., Fleming, L. E. & Osborne, N. J. Prevalence, Associated Factors, and Comorbid Conditions for Ménière’s Disease. Ear Hear. 35, e162 (2014).

7. Harris, J. P. & Alexander, T. H. Current-Day Prevalence of Ménière’s Syndrome. Audiol. Neurotol. 15, 318– 322 (2010).

8. Havia, M., Kentala, E. & PyykkÖ, I. Prevalence of Menière’s Disease in General Population of Southern Finland. Otolaryngol. Neck Surg. 133, 762–768 (2005).

9. Ohmen, J. D. et al. Genetic Evidence for an Ethnic Diversity in the Susceptibility to Ménière’s Disease. Otol. Neurotol. 34, 1336 (2013).

10. Yang, T.-H. et al. Peripheral Vestibular Disorders: Nationwide Evidence From Taiwan. The Laryngoscope 131, 639–643 (2021).

11. Parra-Perez, A. M. & Lopez-Escamez, J.A. Types of Inheritance and Genes Associated with Familial Meniere Disease. J. Assoc. Res. Otolaryngol. 24, 269–279 (2023).

12. Gallego-Martinez, A., Requena, T., Roman-Naranjo, P. & Lopez-Escamez, J. A. Excess of Rare Missense Variants in Hearing Loss Genes in Sporadic Meniere Disease. Front. Genet. 10, (2019).

13. Gallego-Martinez, A., Requena, T., Roman-Naranjo, P., May, P. & Lopez-Escamez, J. A. Enrichment of damaging missense variants in genes related with axonal guidance signalling in sporadic Meniere’s disease. J. Med. Genet. 57, 82–88 (2020).

14. Fisch, K. M. et al. The genomic landscape of Ménière’s disease: a path to endolymphatic hydrops. BMC Genomics 25, 646 (2024).

15. Roman-Naranjo, P. et al. Rare coding variants involving MYO7A and other genes encoding stereocilia link proteins in familial meniere disease. Hear. Res. 409, 108329 (2021).

16. Requena, T. et al. Identification of two novel mutations in FAM136A and DTNA genes in autosomal-dominant familial Meniere’s disease. Hum. Mol. Genet. 24, 1119–1126 (2015).

17. Roman-Naranjo, P. et al. Burden of Rare Variants in the OTOG Gene in Familial Meniere’s Disease. Ear Hear. 41, 1598 (2020).

18. Klockars, T. & Kentala, E. Inheritance of Ménière’s Disease in the Finnish Population. Arch. Otolaryngol. Neck Surg. 133, 73–77 (2007).

19. Morrison, A. W. & Johnson, K. J. Genetics (molecular biology) and Meniere’s disease. Otolaryngol. Clin. North Am. 35, 497–516 (2002).

20. Morrison, A. W., Bailey, M. E. S. & Morrison, G. a. J. Familial Ménière’s disease: clinical and genetic aspects. J. Laryngol. Otol. 123, 29–37 (2009).

21. Escalera-Balsera, A. et al. A rare haplotype of the GJD3 gene segregating in familial Meniere’s disease interferes with connexin assembly. Genome Med. 17, 4 (2025).

22. Uffelmann, E. et al. Genome-wide association studies. Nat. Rev. Methods Primer 1, 1–21 (2021).

23. Bycroft, C. et al. The UK Biobank resource with deep phenotyping and genomic data. Nature 562, 203–209 (2018).

24. The All of Us Research Program Genomics Investigators. Genomic data in the All of Us Research Program. Nature 627, 340–346 (2024).

25. Karczewski, K. J. et al. Pan-UK Biobank genome-wide association analyses enhance discovery and resolution of ancestry-enriched effects. Nat. Genet. 57, 2408–2417 (2025).

26. Kurki, M. I. et al. FinnGen provides genetic insights from a well-phenotyped isolated population. Nature 613, 508–518 (2023).

27. Verma, A. et al. Diversity and scale: Genetic architecture of 2068 traits in the VA Million Veteran Program. Science 385, eadj1182 (2024).

28. Nagai, A. et al. Overview of the BioBank Japan Project: Study design and profile. J. Epidemiol. 27, S2–S8 (2017).

29. Zeggini, E. & Ioannidis, J. P. Meta-Analysis in Genome-Wide Association Studies. Pharmacogenomics 10, 191–201 (2009).

30. Evangelou, E. & Ioannidis, J. P. A. Meta-analysis methods for genome-wide association studies and beyond. Nat. Rev. Genet. 14, 379–389 (2013).

31. Li, X. et al. Powerful, scalable and resource-efficient meta-analysis of rare variant associations in large whole genome sequencing studies. Nat. Genet. 55, 154–164 (2023).

32. Mbatchou, J. et al. Computationally efficient whole-genome regression for quantitative and binary traits. Nat. Genet. 53, 1097–1103 (2021).

33. Sakaue, S. et al. A cross-population atlas of genetic associations for 220 human phenotypes. Nat. Genet. 53, 1415–1424 (2021).

34. Willer, C. J., Li, Y. & Abecasis, G. R. METAL: fast and efficient meta-analysis of genomewide association scans. Bioinformatics 26, 2190–2191 (2010).

35. Bulik-Sullivan, B. K. et al. LD Score regression distinguishes confounding from polygenicity in genome-wide association studies. Nat. Genet. 47, 291–295 (2015).

36. Abe, S., Takeda, H., Nishio, S. & Usami, S. Sensorineural hearing loss and mild cardiac phenotype caused by an EYA4 mutation. Hum. Genome Var. 5, 1–4 (2018).

37. Shinagawa, J. et al. Prevalence and clinical features of hearing loss caused by EYA4 variants. Sci. Rep. 10, 3662 (2020).

38. Depreux, F. F. S. et al. Eya4-deficient mice are a model for heritable otitis media. J. Clin. Invest. 118, 651– 658 (2008).

39. Wang, L. et al. Eya4 regulation of Na+/K+-ATPase is required for sensory system development in zebrafish. Development 135, 3425–3434 (2008).

40. Wayne, S. et al. Mutations in the transcriptional activator EYA4 cause late-onset deafness at the DFNA10 locus. Hum. Mol. Genet. 10, 195–200 (2001).

41. Zou, Y., Carbonetto, P., Wang, G. & Stephens, M. Fine-mapping from summary data with the “Sum of Single Effects” model. PLOS Genet. 18, e1010299 (2022).

42. Wells, H. R. R. et al. GWAS Identifies 44 Independent Associated Genomic Loci for Self-Reported Adult Hearing Difficulty in UK Biobank. Am. J. Hum. Genet. 105, 788–802 (2019).

43. Chang, E. H. et al. Branchio-oto-renal syndrome: The mutation spectrum in EYA1 and its phenotypic consequences. Hum. Mutat. 23, 582–589 (2004).

44. Xu, P.-X. et al. Eya1-deficient mice lack ears and kidneys and show abnormal apoptosis of organ primordia. Nat. Genet. 23, 113–117 (1999).

45. Abdelhak, S. et al. A human homologue of the Drosophila eyes absent gene underlies Branchio-Oto-Renal (BOR) syndrome and identifies a novel gene family. Nat. Genet. 15, 157–164 (1997).

46. Zou, D. et al. Eya1 gene dosage critically affects the development of sensory epithelia in the mammalian inner ear. Hum. Mol. Genet. 17, 3340–3356 (2008).

47. Deng, M., Pan, L., Xie, X. & Gan, L. Requirement for Lmo4 in the vestibular morphogenesis of mouse inner ear. Dev. Biol. 338, 38–49 (2010).

48. Deng, M. et al. LMO4 Functions As a Negative Regulator of Sensory Organ Formation in the Mammalian Cochlea. J. Neurosci. 34, 10072–10077 (2014).

49. Sakai, Y., Luo, T., McCaffery, P., Hamada, H. & Dräger, U. C. CYP26A1 and CYP26C1 cooperate in degrading retinoic acid within the equatorial retina during later eye development. Dev. Biol. 276, 143–157 (2004).

50. White, R. J. & Schilling, T. F. How degrading: Cyp26s in hindbrain development. Dev. Dyn. Off. Publ. Am. Assoc. Anat. 237, 2775–2790 (2008).

51. Emoto, Y., Wada, H., Okamoto, H., Kudo, A. & Imai, Y. Retinoic acid-metabolizing enzyme Cyp26a1 is essential for determining territories of hindbrain and spinal cord in zebrafish. Dev. Biol. 278, 415–427 (2005).

52. Abu-Abed, S. et al. The retinoic acid-metabolizing enzyme, CYP26A1, is essential for normal hindbrain patterning, vertebral identity, and development of posterior structures. Genes Dev. 15, 226–240 (2001).

53. Ono, K. et al. Retinoic acid degradation shapes zonal development of vestibular organs and sensitivity to transient linear accelerations. Nat. Commun. 11, 63 (2020).

54. Moodley, A. & Van Der Meyden, K. Could there be any merit in lumping primary open-angle glaucoma, idiopathic intracranial hypertension and Meniere’s disease into a novel and discrete category of fluid tension disorders? Med. Hypotheses 132, 109361 (2019).

55. Kim, H. et al. The Retinoic Acid Synthesis Gene ALDH1a2 Is a Candidate Tumor Suppressor in Prostate Cancer. Cancer Res. 65, 8118–8124 (2005).

56. Touma, S. E., Perner, S., Rubin, M. A., Nanus, D. M. & Gudas, L. J. Retinoid metabolism and ALDH1A2 (RALDH2) expression are altered in the transgenic adenocarcinoma mouse prostate model. Biochem. Pharmacol. 78, 1127–1138 (2009).

57. El Kares, R. et al. A human ALDH1A2 gene variant is associated with increased newborn kidney size and serum retinoic acid. Kidney Int. 78, 96–102 (2010).

58. Beecroft, S. J. et al. Biallelic hypomorphic variants in ALDH1A2 cause a novel lethal human multiple congenital anomaly syndrome encompassing diaphragmatic, pulmonary, and cardiovascular defects. Hum. Mutat. 42, 506–519 (2021).

59. Ge, Y.-J. et al. Genetic architectures of cerebral ventricles and their overlap with neuropsychiatric traits. Nat. Hum. Behav. 8, 164–180 (2024).

60. Valk, W. H. van der et al. A single-cell level comparison of human inner ear organoids with the human cochlea and vestibular organs. Cell Rep. 42, (2023).

61. Orvis, J. et al. gEAR: Gene Expression Analysis Resource portal for community-driven, multi-omic data exploration. Nat. Methods 18, 843–844 (2021).

62. Chari, D. A. et al. A modern conceptual framework for study and treatment of Meniere’s disease. Front. Neurol. 16, (2025).

63. Romand, R. et al. The retinoic acid receptors RARα and RARγ are required for inner ear development. Mech. Dev. 119, 213–223 (2002).

64. Bok, J. et al. Transient retinoic acid signaling confers anterior-posterior polarity to the inner ear. Proc. Natl. Acad. Sci. 108, 161–166 (2011).

65. Frenz, D. A. et al. Retinoid signaling in inner ear development: A “Goldilocks” phenomenon. Am. J. Med. Genet. A. 152A, 2947–2961 (2010).

66. Maier, E. C. & Whitfield, T. T. RA and FGF Signalling Are Required in the Zebrafish Otic Vesicle to Pattern and Maintain Ventral Otic Identities. PLOS Genet. 10, e1004858 (2014).

67. Kim, S. Y. et al. Association Between Meniere Disease and Migraine. JAMA Otolaryngol. Neck Surg. 148, 457–464 (2022).

68. Hautakangas, H. et al. Genome-wide analysis of 102,084 migraine cases identifies 123 risk loci and subtype-specific risk alleles. Nat. Genet. 54, 152–160 (2022).

69. Skuladottir, A. T. et al. A genome-wide meta-analysis uncovers six sequence variants conferring risk of vertigo. Commun. Biol. 4, 1148 (2021).

70. Gaziano, J. M. et al. Million Veteran Program: A mega-biobank to study genetic influences on health and disease. J. Clin. Epidemiol. 70, 214–223 (2016).

71. Zhou, W. et al. SAIGE-GENE+ improves the efficiency and accuracy of set-based rare variant association tests. Nat. Genet. 54, 1466–1469 (2022).

72. Sakaue, S. et al. A cross-population atlas of genetic associations for 220 human phenotypes. Nat. Genet. 53, 1415–1424 (2021).

73. Perez, G. et al. The UCSC Genome Browser database: 2025 update. Nucleic Acids Res. 53, D1243– D1249 (2025).

74. Auton, A. et al. A global reference for human genetic variation. Nature 526, 68–74 (2015).

75. Lee, S. H., Yang, J., Goddard, M. E., Visscher, P. M. & Wray, N. R. Estimation of pleiotropy between complex diseases using single-nucleotide polymorphism-derived genomic relationships and restricted maximum likelihood. Bioinformatics 28, 2540–2542 (2012).

76. Ojavee, S. E., Kutalik, Z. & Robinson, M. R. Liability-scale heritability estimation for biobank studies of low-prevalence disease. Am. J. Hum. Genet. 109, 2009–2017 (2022).

77. Purcell, S. et al. PLINK: A Tool Set for Whole-Genome Association and Population-Based Linkage Analyses. Am. J. Hum. Genet. 81, 559–575 (2007).

78. McLaren, W. et al. The Ensembl Variant Effect Predictor. Genome Biol. 17, 122 (2016).

79. THE GTEX CONSORTIUM. The GTEx Consortium atlas of genetic regulatory effects across human tissues. Science 369, 1318–1330 (2020).

80. Leeuw, C. A. de, Mooij, J. M., Heskes, T. & Posthuma, D. MAGMA: Generalized Gene-Set Analysis of GWAS Data. PLOS Comput. Biol. 11, e1004219 (2015).

