## Supplemental for "Genome-wide analysis implicates inner ear development in Ménière’s disease": Supplementary_Figures.pdf

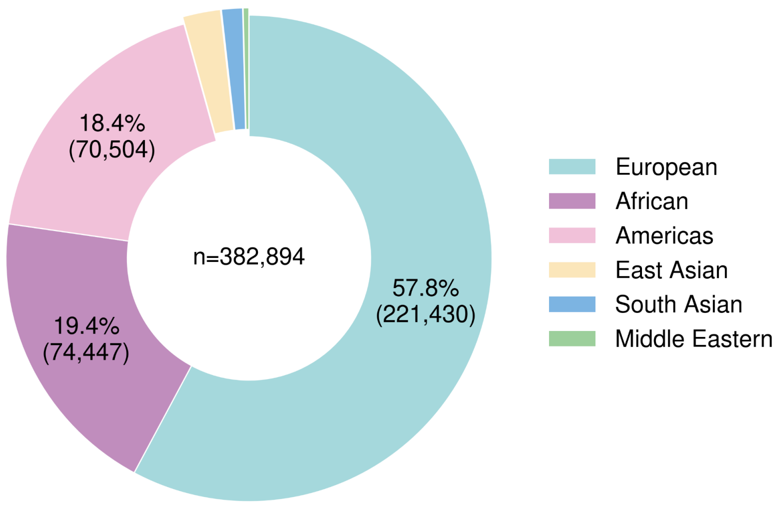


**Supplementary Figure 1.** Genetically inferred ancestry distribution of all individuals in the All of Us version 8.

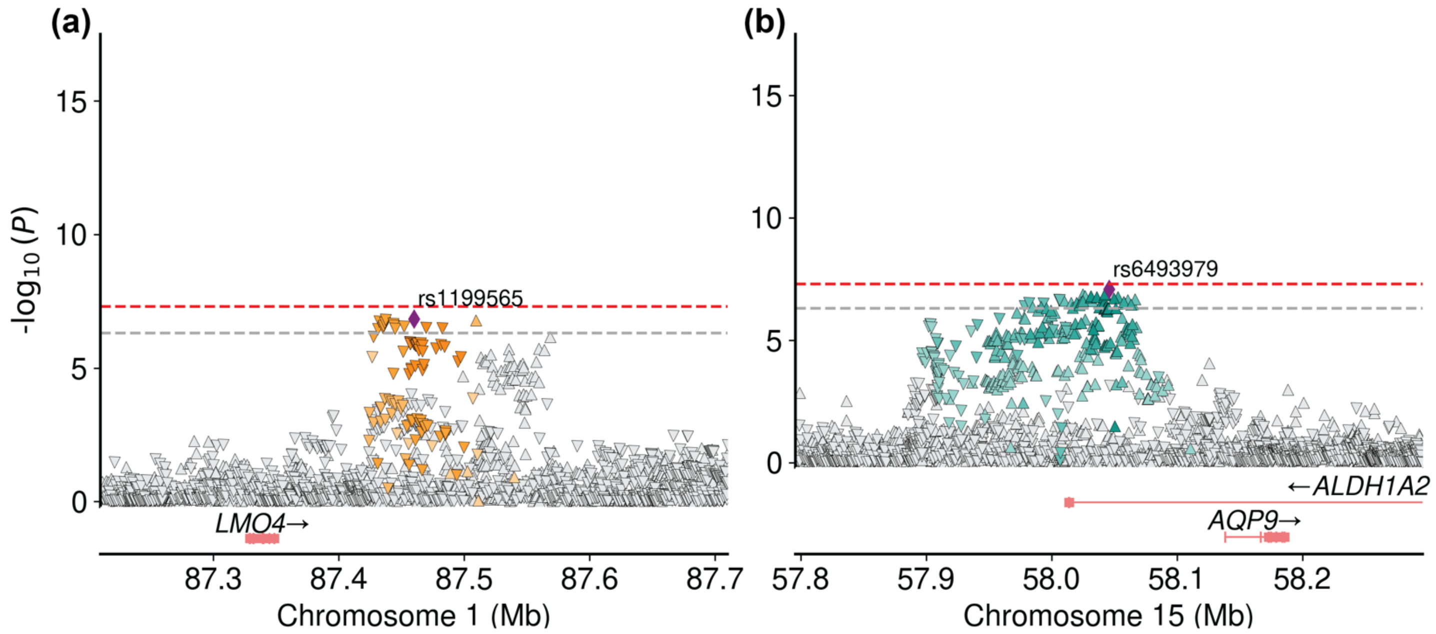


**Supplementary Figure 2. LocusZoom plot of suggestive loci for Ménière's disease.** Negative $\log_{10}$ P-values from the meta-analysis plotted against genomic positions for each index SNP region. The window size of the plots is 500kb. Genes of interest are colored in red. Red dotted line denotes genome-wide significance ($P<5\times10^{-8}$). Grey dotted line denotes suggestive significance ($5\times10^{-8}<P<5\times10^{-7}$).

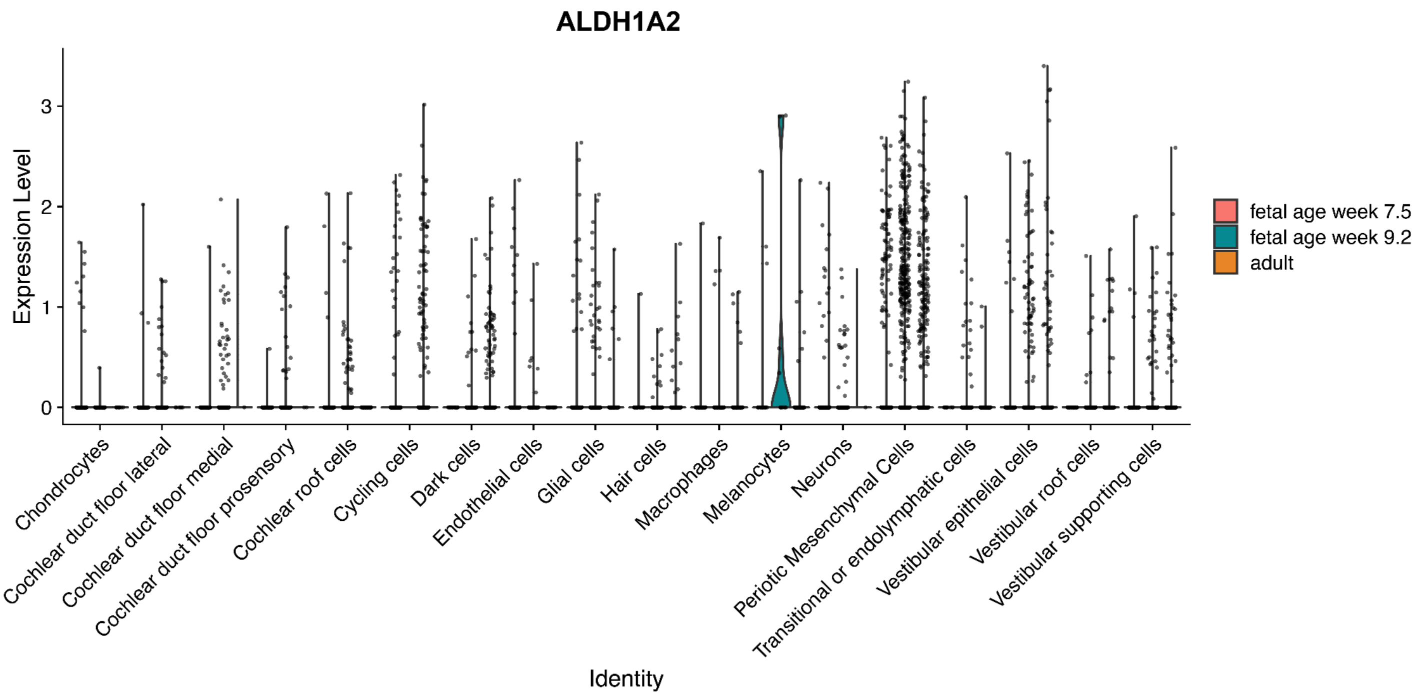


**Supplementary Figure 3. Violin plot of single-cell expression in ALDH1A2 from human inner ear scRNAseq across developmental stages.** *A violin plot shows the expression of ALDH1A2 across diverse human inner ear cell types at three developmental stages (fetal week 7.5, fetal week 9.2, and adult). Data were obtained from the human inner ear atlas*[60] *via the gEAR Dashboard*[61].
